# Investigating the relationship between breast cancer risk factors and an AI-generated mammographic texture feature in the Nurses’ Health Study II

**DOI:** 10.1101/2025.02.18.25322419

**Authors:** Xueyao Wu, Shu Jiang, Aaron Ge, Constance Turman, Graham Colditz, Rulla M. Tamimi, Peter Kraft

## Abstract

**Introduction:** The mammogram risk score (MRS), an AI-driven texture feature derived from digital mammograms, strongly predicts breast cancer risk independently of breast density, though underlying mechanisms remain unclear. This study investigated relationships between established breast cancer risk factors, covering anthropometrics, reproductive factors, family history, and mammographic density metrics, and MRS.

**Methods:** Using data from the Nurses’ Health Study II (292 cases, 561 controls), we validated MRS’s association with breast cancer using logistic regression and evaluated its relationships with risk factors through: linear regressions of MRS on observed risk factors and polygenic scores associated with risk factors, and Mendelian randomization (MR) analysis via two-stage least squares regression. We conducted two-sample MR of MRS using summary statistics from genome-wide association studies of risk factors.

**Results:** MRS was significantly associated with breast cancer risk before adjustment for BI-RADS density (OR=1.92 per SD increase in MRS; 95%CI:1.57-2.33; AUC=0.69) and after (OR=1.85; 95%CI:1.49-2.30). Early life body size and adult body mass index (BMI) were inversely associated with MRS, while history of benign breast disease and BI-RADS density showed positive associations; after adjusting for BI-RADS density, associations between MRS and the other three risk factors attenuated. Higher polygenic score for dense area was associated with increased MRS (β=0.16 SD increase in MRS per SD increase in polygenic score; 95%CI: 0.06-0.25), as was percent density (β=0.14; 95%CI:0.05-0.23). Two-sample MR identified associations between genetically predicted dense area (β=0.83 SD increase in MRS per SD increase in dense area; 95%CI:0.39-1.27) and percent density (β=1.14; 95%CI:0.55-1.74) with MRS. After adjusting for BI-RADS density and BMI, higher waist-to-hip ratio was significantly associated with increased MRS in polygenic score and two-sample MR analyses. No significant associations were observed with other risk factors.

**Conclusion:** We validated MRS’s association with breast cancer risk in cases diagnosed 0.5-10.1 years (median 2.6) after mammogram acquisition. Our findings reveal robust associations between breast density measures and MRS and suggest a potential impact of central obesity on MRS. Future larger-scale studies are crucial to validate these results and explore their potential to enhance our understanding of breast cancer etiology and refine risk prediction models.

## Background

Breast cancer remains the most prevalent malignant cancer among women worldwide^1^. While advances in mammographic screening have facilitated early detection and risk stratification by assessing breast density^2^, traditional measures of mammographic density primarily evaluate the relative amounts of fibroglandular tissue (i.e., the functional breast tissue composed of epithelial and stromal cells). This approach limits our ability to fully capture the heterogeneity of individual breast tissue features, such as architecture and spatial relations^3^. Recent research has leveraged accumulating digital mammogram datasets coupled with sophisticated computational techniques to quantify texture features of mammogram, aiming for more precise and individualized risk predictions. These texture features capture detailed patterns and variations in breast tissue that go beyond simple density measurements^4^. A notable development in this area is the mammogram risk score (MRS), an innovative, artificial intelligence (AI)-driven texture feature derived from whole mammogram images that robustly predicts breast cancer risk independently of breast density (5-year area under the receiver operating characteristic curve [AUC] = 0.75)^5,6^. However, the biological underpinnings of this texture feature remain unclear.

The risk of breast cancer is influenced by multiple factors beyond age and genetic markers. Lifestyle, behavioral, and developmental factors, such as anthropometric measures and reproductive events, collectively contribute to breast cancer susceptibility^7^ and may also relate to features in breast tissue. Epidemiological studies have highlighted significant associations between traditional measures of mammographic density and various risk factors, including early life and adult adiposity^8–11^, height^12,13^, age at menarche^12,13^, age at first birth^14,15^, age at natural menopause^16^, and other reproductive/hormonal factors^17^. Utilizing germline genetic variants as instrumental variables (IVs) to strengthen causal inference, Mendelian randomization (MR) studies have reinforced associations with early life and adult adiposity^18,19^, offering protection against confounding and reverse causation typical in observational studies^20^.

Given that texture features capture distinct aspects of breast tissue than summary density measures, investigating how established risk factors relate to these features could improve our understanding of their underlying biology and provide valuable insights into breast cancer pathogenesis. Previous studies have demonstrated phenotypic and genetic relationships between adiposity and V^21,22^, a texture feature reflecting grayscale intensity variations on digitized film mammograms^23^. Representing an advancement over V, MRS was developed using supervised machine learning to not only predict more accurately variation in whole digital images but also to capture biological features relevant to breast cancer risk^6^. These characteristics make MRS a promising target for investigation aimed at advancing breast cancer prevention. Yet, to date, no observational or MR study has explored these associations for the MRS.

With an overarching goal of deepening the understanding of the biological underpinnings of MRS and its potential role in breast cancer susceptibility, the present study comprehensively investigates the relationships between established breast cancer risk factors - encompassing anthropometrics, reproductive and hormonal factors, family history, and traditional mammographic density metrics - and MRS, through comprehensive observational and genetic analyses performed within the Nurses’ Health Study II (NHS II).

## Methods

### Study participants

The current study leverages resources from the NHS II, a large prospective cohort established in 1989 with 116,429 female and predominantly White (> 90%) registered nurses aged 25 to 42 from 14 states^24^. Between 1996 and 1999, blood samples were collected from 29,611 women, forming a blood subcohort^25^. Genotype data from four platforms (Affymetrix 6.0, Illumina HumanHap, Illumina OmniExpress, and Illumina OncoArray) imputed to the 1000 Genomes Phase 3 version 5 reference panel were used in this study. Pre-diagnostic screening mammograms, conducted as close as possible to the blood draw date, were collected as part of breast cancer case-control study nested within the blood subcohort. Participants have been followed up biennially through self-administered questionnaires to update exposure information and disease diagnoses. For this study, we initially included 853 women (292 cases and 561 controls) with eligible full-field digital mammograms. Among these, 383 women (143 cases and 240 controls) had available imputed genotype data and were included in the genetic analyses. Detailed descriptions of the full genotyping and quality control pipeline^26^, as well as the mammogram collection and processing procedure^21,27^, are available in previous publications. The study protocol was approved by the institutional review boards of the Brigham and Women’s Hospital and Harvard T.H. Chan School of Public Health, and those of participating registries as required.

### Risk factors measurement

Information on various established risk factors for breast cancer was collected for NHS II women. These factors included early life and adult body size, fat distribution, height, reproductive characteristics, and family history of breast cancer. Early life body size, waist-to-hip ratio (WHR), height (inch), and age at menarche were reported via the baseline questionnaire in 1989. Body sizes at ages 5 and 10 years were recalled using Stunkard’s nine-level pictogram (levels 1 to 9: most lean to most overweight)^28^. The average of these two measurements was used to represent early life body size. For other covariates, we used the most recent information from the biennial questionnaires preceding the date of the mammogram. These covariates included: body mass index (BMI), age at first birth, menopausal status, age at natural menopause, current postmenopausal hormone use, parity (number of pregnancies ≥ 6 months), history of benign breast disease, and family history of breast cancer. BMI (kg/m^2^) was calculated by dividing weight (kg) by the square of baseline height (m). WHR adjusted for BMI (WHRadjBMI) was further calculated by regressing WHR on BMI and using the residuals from this regression.

We also assessed predicted Breast Imaging Reporting and Data System (BI-RADS) density using a deep learning algorithm which was previously developed to predict mammographic breast density from digital mammograms^29^. The algorithm categorizes breasts from a (almost entirely fatty) to d (extremely dense), matching an experienced mammographer’s evaluation. We coded these categories as 1, 2, 3, 4, with higher numbers indicating denser breasts. The digital mammograms used for MRS calculation were used to assess predicted BI-RADS density.

### Mammogram risk score measurement

The MRS is an AI-derived score capturing the texture information embedded in the whole digital mammograms, represented by millions of pixels^5,6^. It was developed utilizing 220,868 mammograms from 10,126 racially diverse, initially cancer-free women in the Joanne Knight Breast Health Cohort at Washington University (WashU cohort)^30^, of whom 505 developed breast cancer during follow-up. Validation was performed using 150,352 mammograms from 15,885 women in the Emory Breast Imaging Dataset (EMBED), demonstrating consistently robust predictive performance (5-year AUC = 0.74)^6^. The algorithm, previously described in detail^6^, takes all standard mammogram views (craniocaudal [CC] and/or mediolateral oblique [MLO]) from both breasts as input with the option of additional clinical risk factors. The outputs of the algorithm include MRS which is a transparent weighted sum of feature coefficients, probability of 5-year breast cancer onset, and relative risk for each woman that can be used for risk calibration. For the current study, we applied the algorithm to 1706 full-field digital CC view mammograms from the 853 NHS II women. We used the earliest mammograms available for each woman. If the quality of the initial mammogram was unsatisfactory (e.g., due to being a digitized film), we opted for the next available digital mammogram.

### Genetic variants and polygenic scores for risk factors

We identified the largest available genome-wide association studies (GWAS) conducted among women of European ancestry for early life and adult body size, WHR, WHRadjBMI, height, age at menarche, age at first birth, age at natural menopause, number of children ever born, dense area, non-dense area, and percent density. For each risk factor, we collected lists of genetic variants reported as genome-wide significant (*P* < 5.0 × 10^−8^) in the original female-specific GWAS, along with their beta coefficients. When such variants were not explicitly reported, we applied PLINK’s clumping function^31^ (parameters: *P* < 5.0 × 10^−8^, linkage disequilibrium [LD] r^2^ < 0.001 within a 10 Mb window) to obtain this information. For height, for which no female-specific GWAS is known to be publicly accessible, we used genetic variant information from the largest available sex-combined GWAS. This approach was justified as no statistically significant evidence for sex differences in height genetics has been reported^32^.

From the detected genetic variants, we included those that met the following criteria in the NHS II imputed genotype data: matching alleles, non-ambiguous, minimum imputation score > 0.3 across the four genotyping platforms, and minor allele frequency > 0.005. These selected variants were used for the polygenic score (PGS) calculation and as instrumental variables (IVs) in causal inference analyses. Detailed information on GWAS sources and quality control of genetic variants is provided in **Table S1**.

### Statistical analysis

We generated descriptive statistics for all variables. Continuous variables were described using mean and standard deviation, while categorical variables were described using frequency and percentage. We assessed differences between higher and lower MRS groups (using the median value as the cutoff) using Student’s t-test or Wilcoxon rank-sum test for continuous variables and Chi-square test for categorical variables.

We first validated the association between MRS and breast cancer in NHS II using logistic regression after excluding 91 cases diagnosed within 6 months after the mammogram used for calculation. To evaluate the association between breast cancer risk factors and MRS, four main analyses were performed: (i) linear regressions of MRS on each observed risk factor to quantify their observational association without accounting for genetic predisposition; (ii) linear regressions of MRS on the PGS associated with each risk factor to evaluate the relationship between genetic predisposition to each risk factor and MRS; (iii) Mendelian randomization (MR) analysis via two-stage least squares (2SLS) regressions of MRS on each genetically predicted risk factor, and (iv) two-sample MR of MRS using GWAS summary statistics of each risk factor, to evaluate potential causal associations. For all analyses, we standardized MRS and all non-binary variables for easier comparison across risk factors. Binary variables included postmenopausal hormone use, history of benign breast disease, and family history of breast cancer, each categorized as “Yes” or “No”. In two-sample MR, we retained the original scale of genetic associations from the source GWAS. For each risk factor, we calculated its weighted PGS using PLINK’s “--score” function^31^, summing the products of effect allele dosage and corresponding beta coefficient across all selected genetic variants for each woman. Prior to 2SLS regression, we assessed instrument strength by regressing each risk factor on its corresponding PGS, obtaining *F*-statistics and correlation coefficient estimates. To minimize weak instrument bias, we excluded PGS with *F*-statistic < 5 or correlation *P* > 0.05 from the 2SLS analysis. While we calculated the association between PGS for both dense area and percent density with BI-RADS density, BI-RADS density was excluded from 2SLS analysis given the inconsistent phenotype definitions between BI-RADS and the objective density measurements in GWAS. The 2SLS procedure involved two stages: first, regressing each risk factor on its PGS; second, using the predicted values as independent variables in a regression model with MRS as the dependent variable.

For two-sample MR, we obtained the “IVs-exposure” associations directly from the corresponding GWAS, and used PLINK’s “--glm” function^31^ to obtain association estimates between each genetic variant and MRS in the NHS II dataset, representing the “IVs-outcome” associations. Our primary method was the random-effect inverse-variance weighted (IVW) approach^33^, which assumes a zero intercept and estimates causality using random-effects meta-analysis. To validate MR model assumptions^20^ and assess the robustness of our findings, we applied complementary methods including MR-Egger regression (which detects and accounts for directional pleiotropy)^34^, weighted median (robust to up to 50% invalid instruments)^35^, weighted mode (identifies the causal effect estimate that is most consistent across all variants)^36^, and IVW excluding outlier SNPs detected using Radial MR’s iterative Cochran’s Q method^37^. We considered a causal association significant if it reached statistical significance in the IVW analysis and maintained consistent direction across all sensitivity analyses. Following two-sample MR, we performed two additional analyses: an MR-Clust analysis to cluster genetic variants with similar causal estimates, which may reflect heterogeneous causal mechanisms^38^, and a Chi-square test on Wald ratios estimated in two-sample MR across all IVs for each risk factor to test if any of the risk factor-associated genetic variants associates with MRS.

To mitigate confounding, we employed three adjustment sets across all analyses. The crude model included age at mammogram and, where appropriate, genotyping platform and the top ten genetic principal components. The second and third sets additionally adjusted for menopausal status and predicted BI-RADS density, respectively. To address potential ascertainment bias arising from investigating MRS in a case-control study design that implicitly conditions on breast cancer status, we conducted all analyses using two approaches as previously recommended^39^: (1) including case-control status as an additional covariate (our primary approach to maintain sample size), and (2) restricting analyses to controls only. 2SLS and TSMR analyses were conducted using packages “ivreg”, “TwoSampleMR”, and “RadialMR” in R (v4.1.0). We used the conventional *P*-value threshold of 0.05 to define statistical significance given the relatively limited sample size and the exploratory nature of our study.

## Results

### Participant characteristics and MRS-breast cancer association in the NHS II

This nested case-control study comprised 853 women (292 cases and 561 controls) with mean age 55.3 (± 5.45 years) at the time of mammogram. The majority (64.1%) were postmenopausal. A comparison of risk factors distribution between groups divided by median MRS can be found in **Table 1**. Notably, compared to those below the median, participants with above-median MRS were younger, more likely to have lower BMI and WHR, denser breasts, a history of benign breast disease at the time of mammogram (*P* < 0.001), and more likely to be breast cancer cases (51.0% vs 23.6%, *P* < 0.001).

**Table 1.**
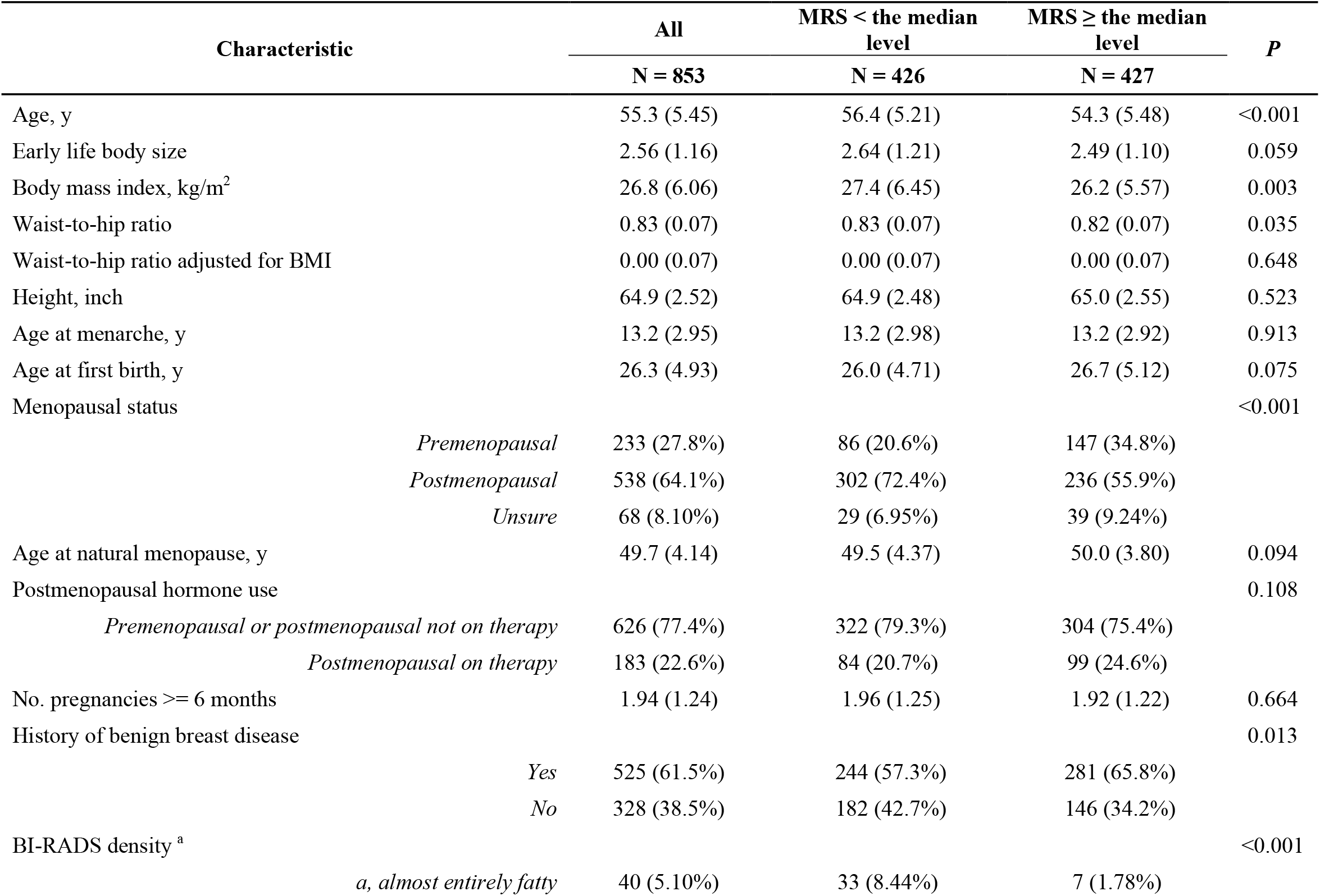
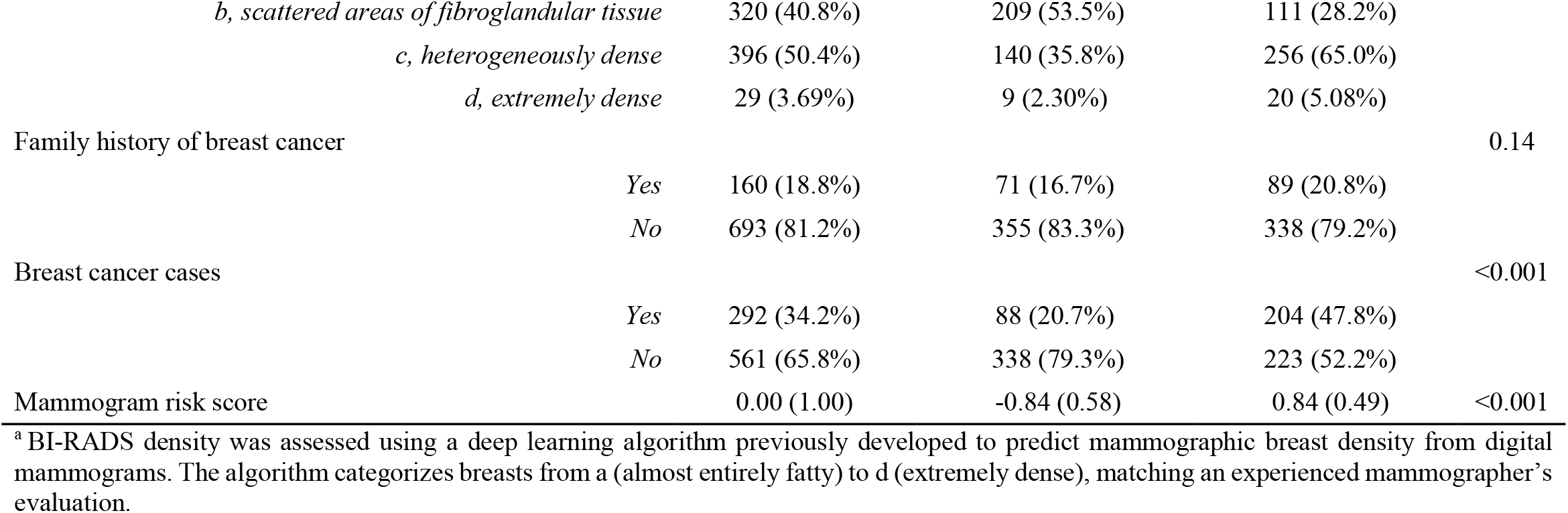
Baseline characteristics of participants according to median mammogram risk score.

External validation in NHS II demonstrated a strong association between MRS and breast cancer risk (odds ratio = 1.92 per standard deviation [SD] difference in MRS; 95% CI: 1.57-2.33; *P* = 1.98×10^−18^; 10-year AUC = 0.69) (**Table S2, Figures S1-S2**). Cases were diagnosed 0.5 to 10.1 years (median 2.6) after the mammogram used for MRS calculation. The association remained unchanged after adjusting for predicted BI-RADS density (**Table S2**).

### Associations between observed breast cancer risk factors and MRS

Both predicted BI-RADS density (β = 0.33 SD difference in MRS per SD difference in BI-RADS density; 95% CI: 0.23-0.42; *P* = 8.75×10^−11^) and history of benign breast disease (β = 0.23 SD difference in MRS with vs. without history; 95% CI: 0.10-0.36; *P* = 4.50×10^−4^) showed positive associations with MRS. Early life body size (β = –0.08 SD difference in MRS per SD difference in body size; 95% CI: –0.15 to –0.02; *P* = 9.59×10^−3^) and adult BMI (β = –0.08 SD difference in MRS per SD difference in BMI; 95% CI: –0.15 to –0.02; *P* = 1.11×10^−2^) demonstrated negative associations. No statistically significant associations were observed for the other examined risk factors (all *P* > 0.05, **Table 2**). These results remained largely consistent when restricted to controls only or additionally adjusted for menopausal status (**Tables S3-S4**). When additionally adjusted for predicted BI-RADS density, associations with history of benign breast disease, early life body size, and adult BMI all attenuated (**Table 2, Table S3**).

**Table 2.**
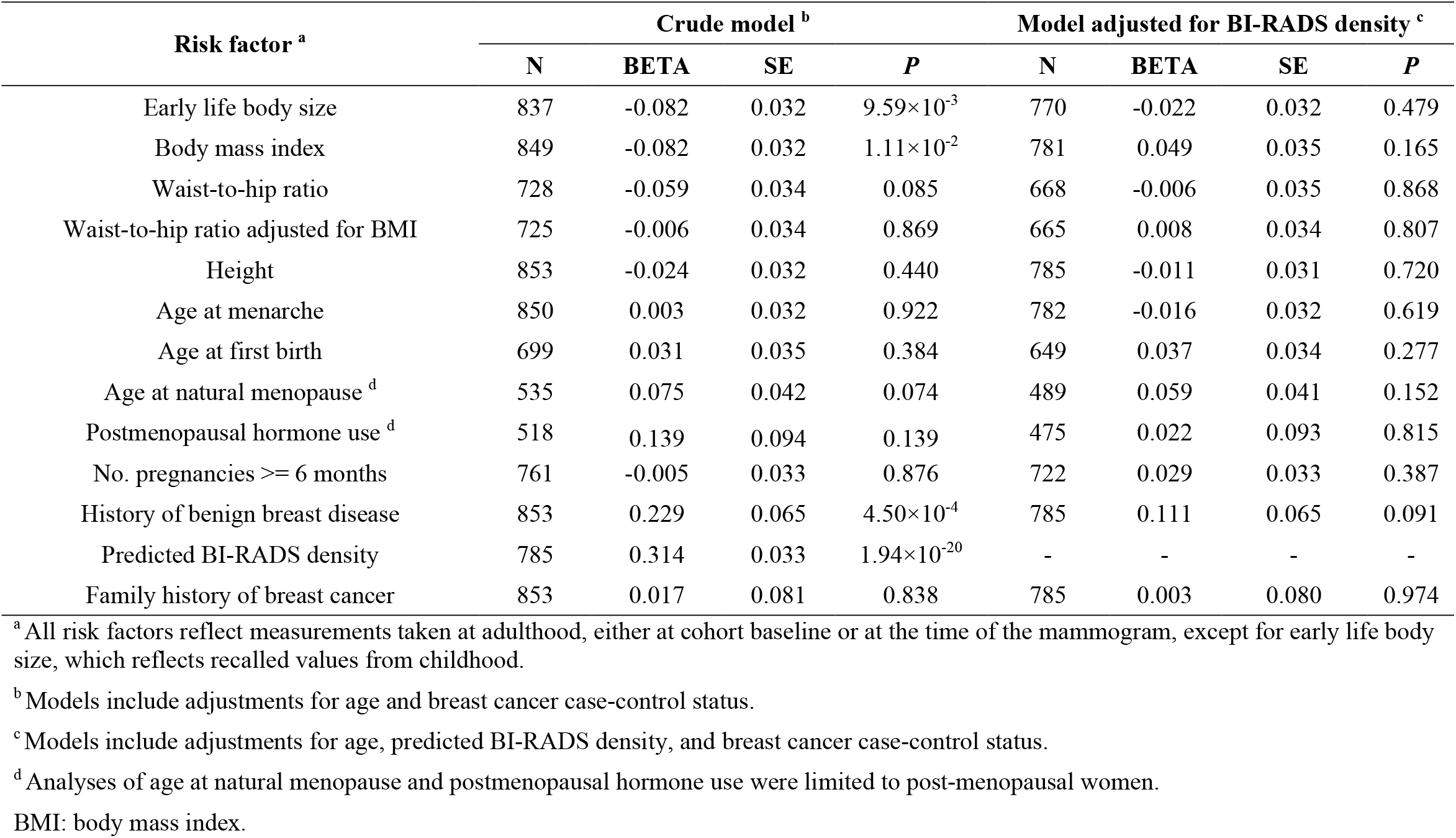
Linear regression of mammogram risk score on each breast cancer risk factor.

### Associations between polygenic scores for breast cancer risk factors and MRS

Linear regressions of MRS on PGS for risk factors revealed significant positive associations for dense area (β = 0.16 SD difference in MRS per SD difference in PGS; 95% CI: 0.06-0.25; *P* = 1.37×10^−3^) and percent density (β = 0.14 SD difference in MRS per SD difference in PGS; 95% CI: 0.05-0.23; *P* = 3.29×10^−3^). No significant associations were observed between the PGS for other risk factors and MRS (**Table 3**). A similar pattern of associations was observed in analyses restricted to controls and in models adjusted for menopausal status (**Tables S5-S6**). After adjusting for predicted BI-RADS density, the association for percent density remained strong, whereas association for dense area weakened slightly. A significant association was additionally observed between higher PGS for WHRadjBMI and increased MRS (β = 0.12 SD difference in MRS per SD difference in PGS; 95% CI: 0.03-0.21; *P* = 1.18×10^−2^) (**Table 3, Table S5**).

**Table 3.**
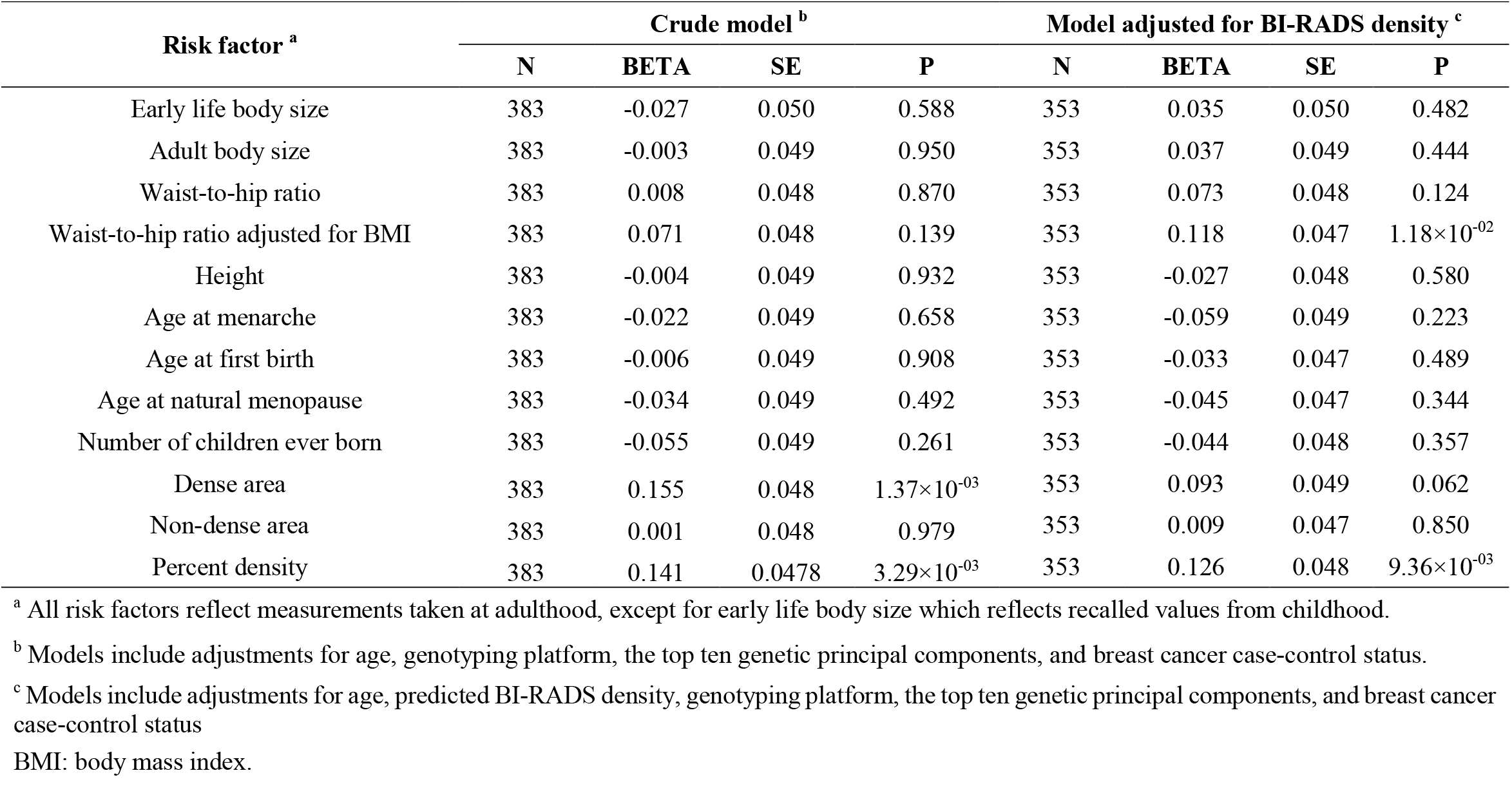
Linear regression of mammogram risk score on polygenic score for each breast cancer risk factor.

| Risk factor <sup>a</sup> | Crude model <sup>b</sup> |  |  |  | Model adjusted for BI-RADS density <sup>c</sup> |  |  |  |
| --- | --- | --- | --- | --- | --- | --- | --- | --- |
|  | N | BETA | SE | P | N | BETA | SE | P |
| Early life body size | 383 | -0.027 | 0.050 | 0.588 | 353 | 0.035 | 0.050 | 0.482 |
| Adult body size | 383 | -0.003 | 0.049 | 0.950 | 353 | 0.037 | 0.049 | 0.444 |
| Waist-to-hip ratio | 383 | 0.008 | 0.048 | 0.870 | 353 | 0.073 | 0.048 | 0.124 |
| Waist-to-hip ratio adjusted for BMI | 383 | 0.071 | 0.048 | 0.139 | 353 | 0.118 | 0.047 | 1.18×10<sup>-02</sup> |
| Height | 383 | -0.004 | 0.049 | 0.932 | 353 | -0.027 | 0.048 | 0.580 |
| Age at menarche | 383 | -0.022 | 0.049 | 0.658 | 353 | -0.059 | 0.049 | 0.223 |
| Age at first birth | 383 | -0.006 | 0.049 | 0.908 | 353 | -0.033 | 0.047 | 0.489 |
| Age at natural menopause | 383 | -0.034 | 0.049 | 0.492 | 353 | -0.045 | 0.047 | 0.344 |
| Number of children ever born | 383 | -0.055 | 0.049 | 0.261 | 353 | -0.044 | 0.048 | 0.357 |
| Dense area | 383 | 0.155 | 0.048 | 1.37×10<sup>-03</sup> | 353 | 0.093 | 0.049 | 0.062 |
| Non-dense area | 383 | 0.001 | 0.048 | 0.979 | 353 | 0.009 | 0.047 | 0.850 |
| Percent density | 383 | 0.141 | 0.0478 | 3.29×10<sup>-03</sup> | 353 | 0.126 | 0.048 | 9.36×10<sup>-03</sup> |

<sup>a</sup> All risk factors reflect measurements taken at adulthood, except for early life body size which reflects recalled values from childhood.
<sup>b</sup> Models include adjustments for age, genotyping platform, the top ten genetic principal components, and breast cancer case-control status.
<sup>c</sup> Models include adjustments for age, predicted BI-RADS density, genotyping platform, the top ten genetic principal components, and breast cancer case-control status
BMI: body mass index.

### Mendelian randomization between breast cancer risk factors and MRS

Linear regressions of risk factors on their corresponding PGS revealed significant genetic associations for 6 risk factors, including height, BMI, age at menarche, early life body size, WHR, and age at natural menopause (*F*-statistics: 212.56 to 5.36, **Table S7**). PGS for dense area was significantly associated with BI-RADS density (R^2^ = 0.03, *F* = 9.09, *P* = 2.76 ×10^−3^), while PGS for percent density showed no association (R^2^ = 0.00, *F* = 1.47, *P* = 0.23). WHRadjBMI, age at first birth, and number of children ever born were excluded from subsequent 2SLS analyses due to weak instrument strength (*F*-statistic < 5). Associations adjusted for menopausal status or BI-RADS density are detailed in **Tables S8-S9**.

Despite these genetic associations, 2SLS analyses identified no statistically significant associations between the 6 genetically predicted risk factors and MRS. The strongest effect estimates were observed for age at natural menopause (β = –0.80 SD difference in MRS per SD difference in genetically predicted age at natural menopause, 95% CI: –2.58-0.98, *P* = 0.38) and early life body size (β = –0.13 SD difference in MRS per SD difference in genetically predicted early life body size, 95% CI: –0.58-0.32, *P* = 0.57), neither of which reached statistical significance (**Table 4**). All additional adjustments yielded similar non-significant results (**Table 4, Tables S10-S11**).

**Table 4.** Two-stage least squares regression between each breast cancer risk factor (exposure) and mammogram risk score (outcome).

| Risk factor <sup>a</sup> | Crude model <sup>b</sup> |  |  |  | Model adjusted for BI-RADS <sup>c</sup> |  |  |  |
| --- | --- | --- | --- | --- | --- | --- | --- | --- |
|  | N | BETA | SE | P | N | BETA | SE | P |
| Early life body size | 370 | -0.132 | 0.234 | 0.574 | 341 | 0.215 | 0.269 | 0.424 |
| Body mass index | 381 | -0.012 | 0.175 | 0.946 | 351 | 0.153 | 0.201 | 0.446 |
| Waist-to-hip ratio | 336 | 0.081 | 0.304 | 0.790 | 307 | 0.702 | 0.542 | 0.196 |
| Height | 383 | -0.007 | 0.084 | 0.932 | 353 | -0.045 | 0.080 | 0.579 |
| Age at menarche | 383 | -0.081 | 0.183 | 0.659 | 353 | -0.222 | 0.186 | 0.232 |
| Age at natural menopause <sup>d</sup> | 259 | -0.800 | 0.907 | 0.378 | 240 | -1.068 | 1.388 | 0.442 |

<sup>a</sup> All risk factors reflect measurements taken at adulthood, except for early life body size which reflects recalled values from childhood.
<sup>b</sup> Models include adjustments for age, genotyping platform, the top ten genetic principal components, and breast cancer case-control status.
<sup>c</sup> Models include adjustments for age, predicted BI-RADS density, genotyping platform, the top ten genetic principal components, and breast cancer case-control status.
<sup>d</sup> Analyses of age at natural menopause were restricted to post-menopausal women.

Two-sample MR analyses identified significant associations between genetically predicted dense area and MRS (β = 0.83 SD difference in MRS per SD difference in dense area; 95% CI: 0.39-1.27; *P* = 2.09×10^−4^), and genetically predicted percent density and MRS (β = 1.14 SD difference in MRS per SD difference in percent density; 95% CI: 0.55-1.74; *P* = 1.61×10^−4^). No evidence supported significant causal associations with other risk factors (**Figure 1**). Sensitivity analyses using MR-Egger regression, weighted median, weighted mode, and IVW excluding outlier SNPs yielded consistent results (**Table S12**). MR-Clust analysis found all variants for dense area and percent density clustered into a single group with similar causal effects, suggesting no evidence of heterogeneous causal mechanisms; for other risk factors, no variants showed significant effects (**Figure S3**). Chi-square tests on Wald ratios across all IVs for each risk factor revealed no statistically significant associations between risk factor-associated genetic variants and MRS (all *P* > 0.05, **Table S13**).

**Figure 1.**
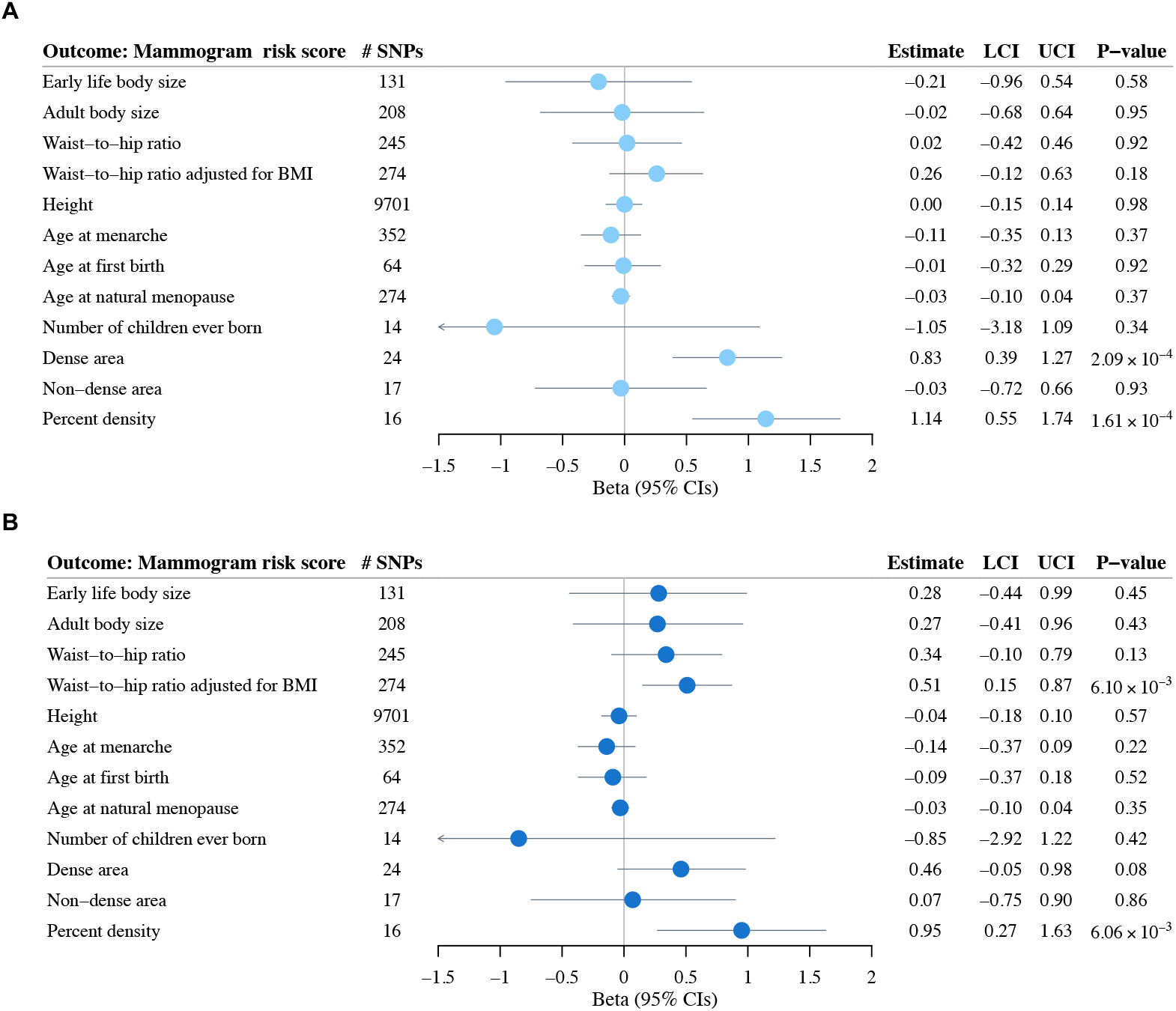
Two-sample Mendelian randomization analysis examining associations between genetically predicted risk factors (exposures) and mammogram risk score (outcome). Results are shown (**A**) before and (**B**) after adjusting for predicted BI-RADS breast density in NHS II. Point estimates (circles) and 95% confidence intervals (error bars) were calculated using the inverse-variance weighted approach. All risk factors represent adult measurements, except for early life body size which was retrospectively reported for childhood. BMI: body mass index; LCI: lower confidence interval; UCI: upper confidence interval.

The patterns of associations were robust to both restriction to control subjects (**Tables S12**) and adjustment for menopausal status (**Tables S14-S15, Figure S4**). Utilizing IV-outcome associations adjusting for predicted BI-RADS density, two-sample MR showed a significant association between genetically predicted WHRadjBMI and MRS (IVW: β = 0.51 SD difference in MRS per SD difference in WHRadjBMI; 95% CI: 0.14-0.88; *P* = 6.10×10^−3^), consistent across all sensitivity analyses. Association with percent density remained substantially unchanged, while association for genetically predicted dense area attenuated (**Tables S16-S17, Figure S5**).

## Discussion

To the best of our knowledge, this study presents one of the first and most comprehensive examinations to date of the relationships between known breast cancer risk factors and MRS - an AI-generated mammographic texture feature derived from full-field digital mammograms. Our analyses revealed robust phenotypic and genetic associations between various mammographic density measures - including predicted BI-RADS density, absolute dense area, and percent density - and the MRS, as well as a suggestive association between higher WHRadjBMI and increasing MRS.

Our external validation of the MRS in the predominantly White NHS II cohort demonstrated robust long-term predictive capability for breast cancer risk. The MRS maintained good discriminatory power (10-year AUC = 0.69) over an extended time horizon, comparing favorably with previous 5-year validations in racially diverse cohorts (WashU: AUC = 0.75; EMBED: AUC = 0.74; 27-46% Non-Hispanic Black women)^6^. Notably, the association between MRS and breast cancer incidence remained robust after adjusting for BI-RADS density, suggesting that MRS provides additional discriminatory power beyond summary density measurements. Conversely, the association between predicted BI-RADS density and breast cancer risk was substantially attenuated after adjusting for MRS. These findings collectively underscore MRS’s generalizability and stability as a risk marker, highlighting its potential to enhance breast cancer risk stratification across diverse clinical settings and populations.

Our findings align with previously reported significant phenotypic relationships between mammographic density and breast texture features^17,21,27,40–43^. The moderate correlation between BI-RADS density and MRS (r ~ 0.33) further corroborates that while these measures are related, they are likely to reflect distinct aspects of mammographic information. Moreover, the observed significant associations between genetic predisposition to higher mammographic density and increased MRS further suggest a shared genetic basis and potential causal relationship underlying these features. This genetic connection enhances the credibility of MRS as a biologically plausible risk factor for breast cancer. Future studies should aim to elucidate the specific biological processes reflected by MRS and their implications for breast cancer etiology.

Investigating the effects of lifestyle, behavioral, and developmental/biological risk factors on breast tissue characteristics, as summarized in mammograms, is crucial for extracting biological insights into modifiable factors for prevention studies and understanding pathways for potential preventive drug targets. While MRS itself represents a novel feature with limited existing literature, it is important to contextualize our findings within the broader landscape of mammographic feature research. Previous studies have demonstrated associations between various breast cancer risk factors and mammographic density^8–16,18,19^, as well as with other texture features such as V^21,22^. Our study of MRS, which incorporates breast architecture and spatial information from whole, digital mammograms, represents an important advancement in understanding the complex relationships between breast tissue features and cancer susceptibility.

The emergence of a statistically significant association between genetic predictors of WHRadjBMI and MRS only after adjusting for BI-RADS density suggests that fat distribution, independent of overall body mass, might influence breast tissue characteristics in ways not fully captured by mammographic density alone. The ability of MRS to reveal this relationship indicates its value as an advanced imaging feature in reflecting nuanced aspects of breast tissue composition that may be relevant to cancer risk assessment. Future studies are needed to validate our results and investigate the biological mechanisms underlying the complex interplay between fat distribution, breast tissue texture features, and breast cancer susceptibility.

Several limitations of our study should be acknowledged. First, our sample size was relatively limited, which may have led to insufficient statistical power to detect associations with some risk factors, particularly those with smaller effect sizes. We emphasize that null findings observed should not be interpreted as definitive evidence of no association, and that larger studies with greater statistical power are needed. Additionally, our analysis was based on a nested case-control design, which could potentially introduce ascertainment bias. However, we expect that this design would not substantially affect our results, given our careful adjustment for case-control status and the consistency of our findings in control-only analyses^39^. Finally, while we evaluated predicted BI-RADS density (which mimics qualitative visual assessments by radiologists)^29^, we were unable to adjust for quantitative density measures due to data unavailability. This limitation may have reduced our statistical power to detect associations, potentially underestimating relationships between other breast cancer risk factors and MRS.

Our study has several notable strengths. A key advantage is the availability of genetic data, digital mammogram data, and comprehensive covariate data on the same set of samples, allowing for integrated analyses across multiple domains. Triangulating evidence from both observational and genetic studies mitigated biases inherent in each study design, providing a multi-perspective evaluation of associations. The MRS algorithm was developed independently of the NHS II cohort, reducing the possibility for overfitting or circular reasoning in our analyses. Developed using standard digital mammograms, sophisticated statistical methods, and large-scale populations, the MRS itself represents an advanced texture feature with significant potential in clinical settings.

## Conclusion

To conclude, this study provides novel insights into the relationships between established breast cancer risk factors and MRS. Findings underscore MRS’s potential independent contribution to breast cancer risk assessment and reveal a potential role of central adiposity in breast tissue characteristics not fully captured by traditional density measures. Future research should encompass larger-scale studies to definitively assess these associations and explore the underlying biological mechanisms. As our understanding of mammographic texture features advances, MRS and similar tools may become integral to personalized breast cancer risk assessment and prevention strategies, offering nuanced insights beyond conventional measures.

## Acknowledgements

The authors would like to acknowledge the contribution to this study from central cancer registries supported through the Centers for Disease Control and Prevention’s National Program of Cancer Registries (NPCR) and/or the National Cancer Institute’s Surveillance, Epidemiology, and End Results (SEER) Program. Central registries may also be supported by state agencies, universities, and cancer centers. Participating central cancer registries include the following: Alabama, Alaska, Arizona, Arkansas, California, Colorado, Connecticut, Delaware, Florida, Georgia, Hawaii, Idaho, Indiana, Iowa, Kentucky, Louisiana, Massachusetts, Maine, Maryland, Michigan, Mississippi, Montana, Nebraska, Nevada, New Hampshire, New Jersey, New Mexico, New York, North Carolina, North Dakota, Ohio, Oklahoma, Oregon, Pennsylvania, Puerto Rico, Rhode Island, Seattle SEER Registry, South Carolina, Tennessee, Texas, Utah, Virginia, West Virginia, Wyoming.

## Authors’ contribution

PK and XW conceived and designed the study. RMT prepared the mammographic texture variation data for NHS II. CT prepared the genotype data for NHS II. SJ and GC prepared the mammogram risk score for NHS II. XW prepared the risk factors GWAS data. XW analyzed the data with the assistance of AG. XW and PK interpreted the results, with significant inputs and comments from RMT, SJ, and GC. XW was a major contributor in writing the manuscript. All authors read and approved the final manuscript.

## Availability of data and materials

The data that support the findings of this study are available from the Nurses’ Health Studies; however, they are not publicly available. Investigators interested in using the data can request access, and feasibility will be discussed at an investigators’ meeting. Limits are not placed on scientific questions or methods, and there is no requirement for co-authorship. Additional data sharing information and policy details can be accessed at http://www.nurseshealthstudy.org/researchers. All GWAS summary statistics used in this study are publicly available.

