## Supplementary material for "Investigating the relationship between breast cancer risk factors and an AI-generated mammographic texture feature in the Nurses’ Health Study II": Figure S

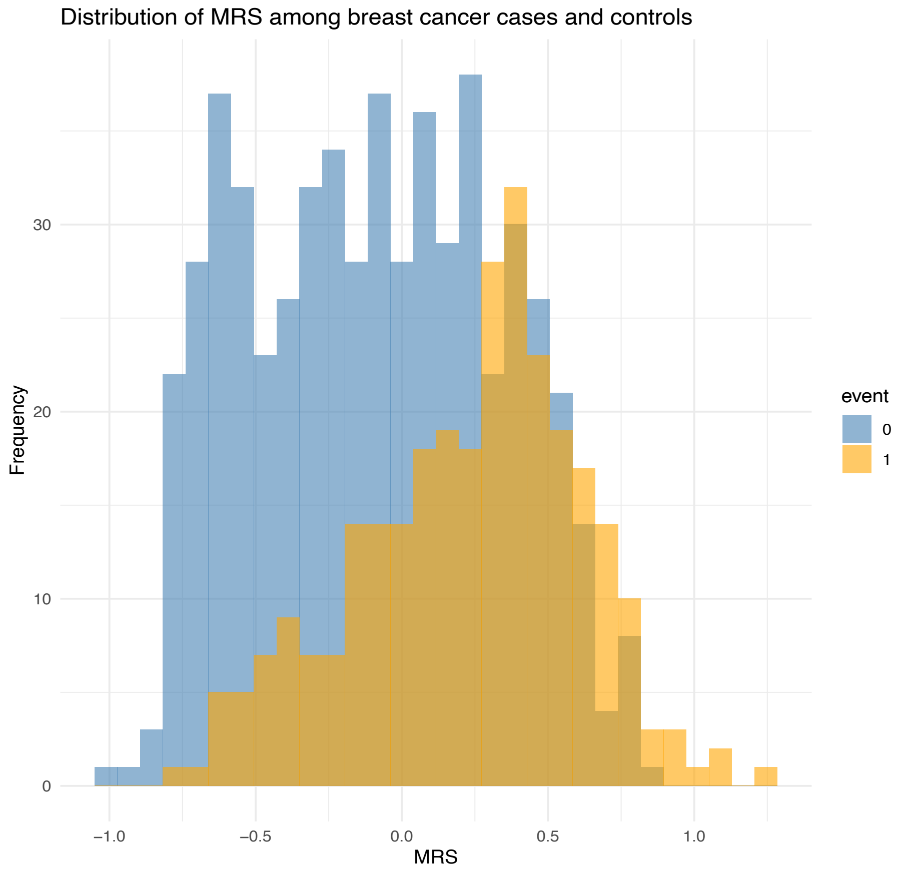

**Figure S1.** Distribution of mammogram risk score (MRS) among breast cancer cases and controls in NHS II.

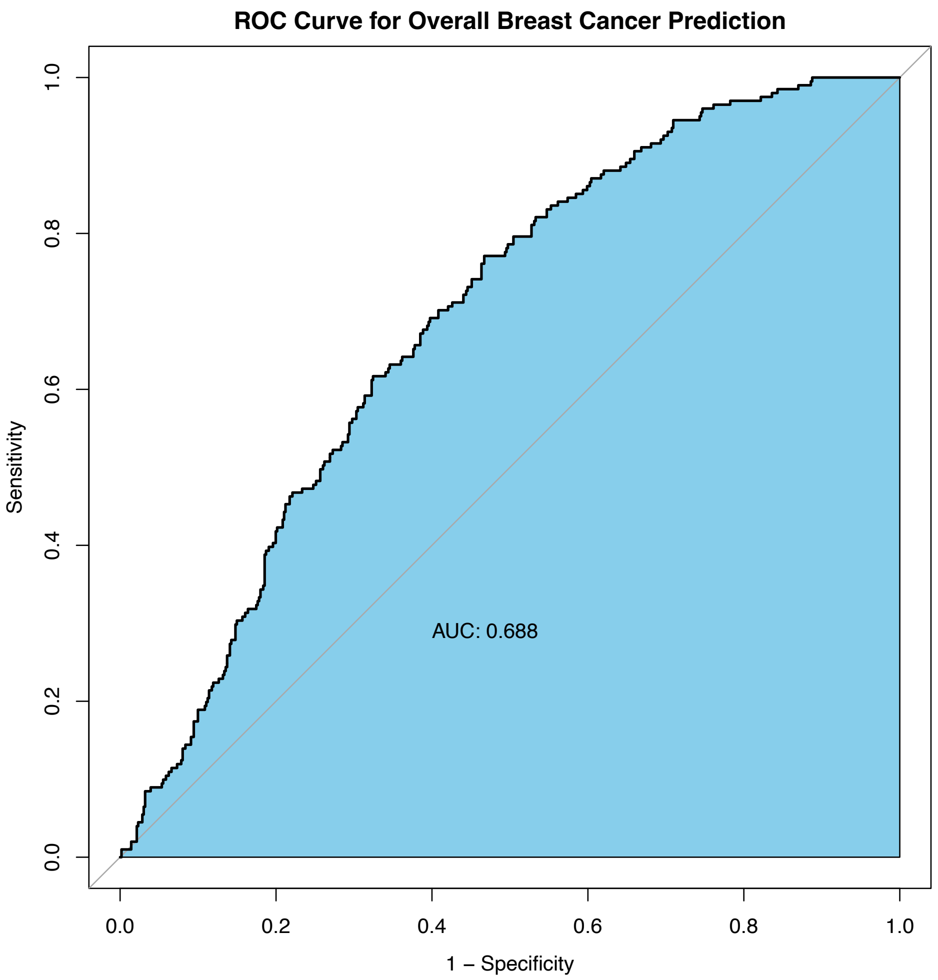


**Figure S2.** Evaluating the performance of mammogram risk score in breast cancer risk prediction in NHS II.
ROC, receiver operating characteristic. AUC, area under the ROC curve.

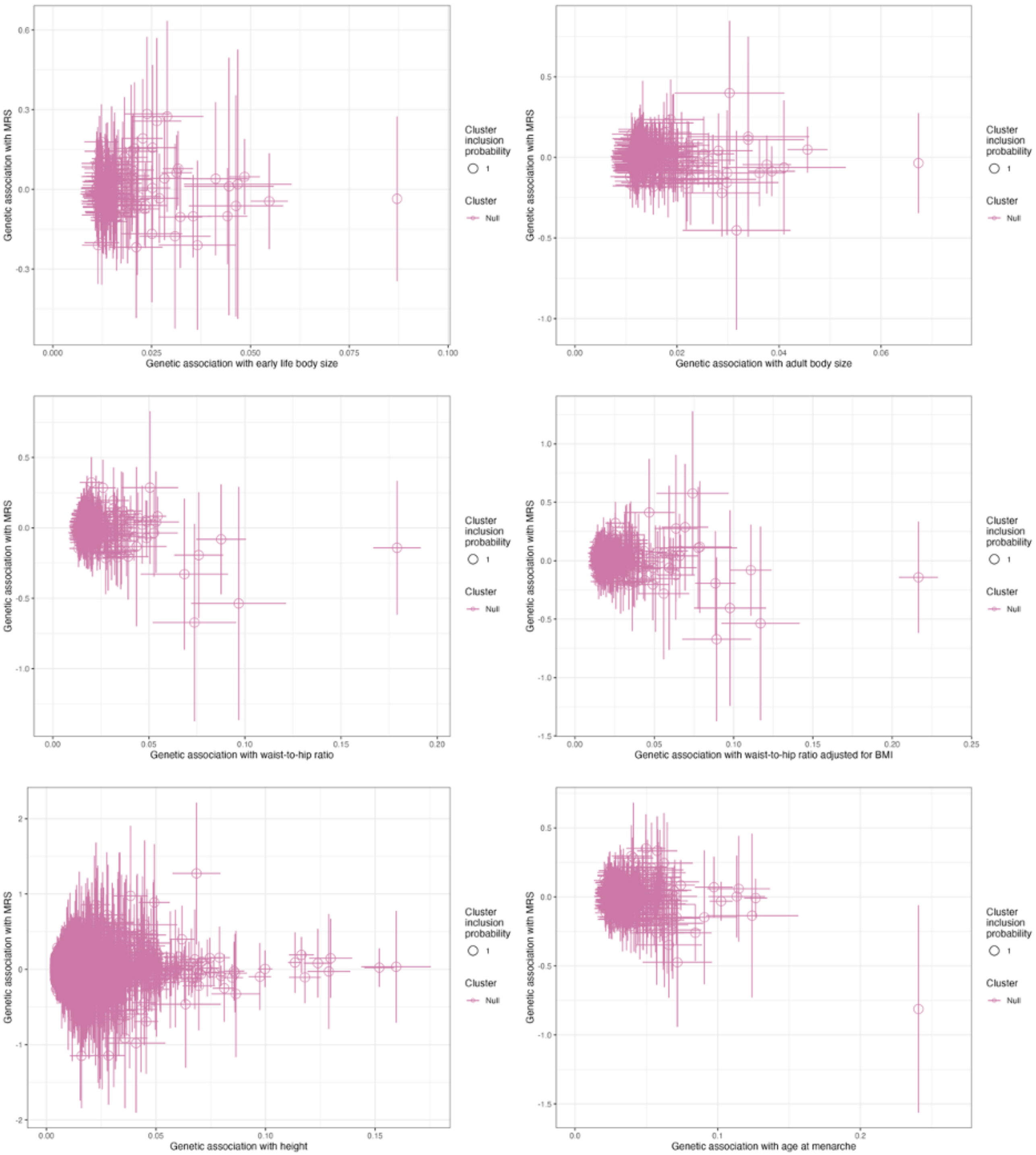


**Figure S3.** Genetic associations between each risk factor and mammogram risk score (MRS). Scatter plots show results from MR-Clust analysis, with genetic association with MRS (y-axis) versus genetic association with each risk factor (x-axis). Each point represents a genetic variant, with lines indicating 95% confidence intervals.

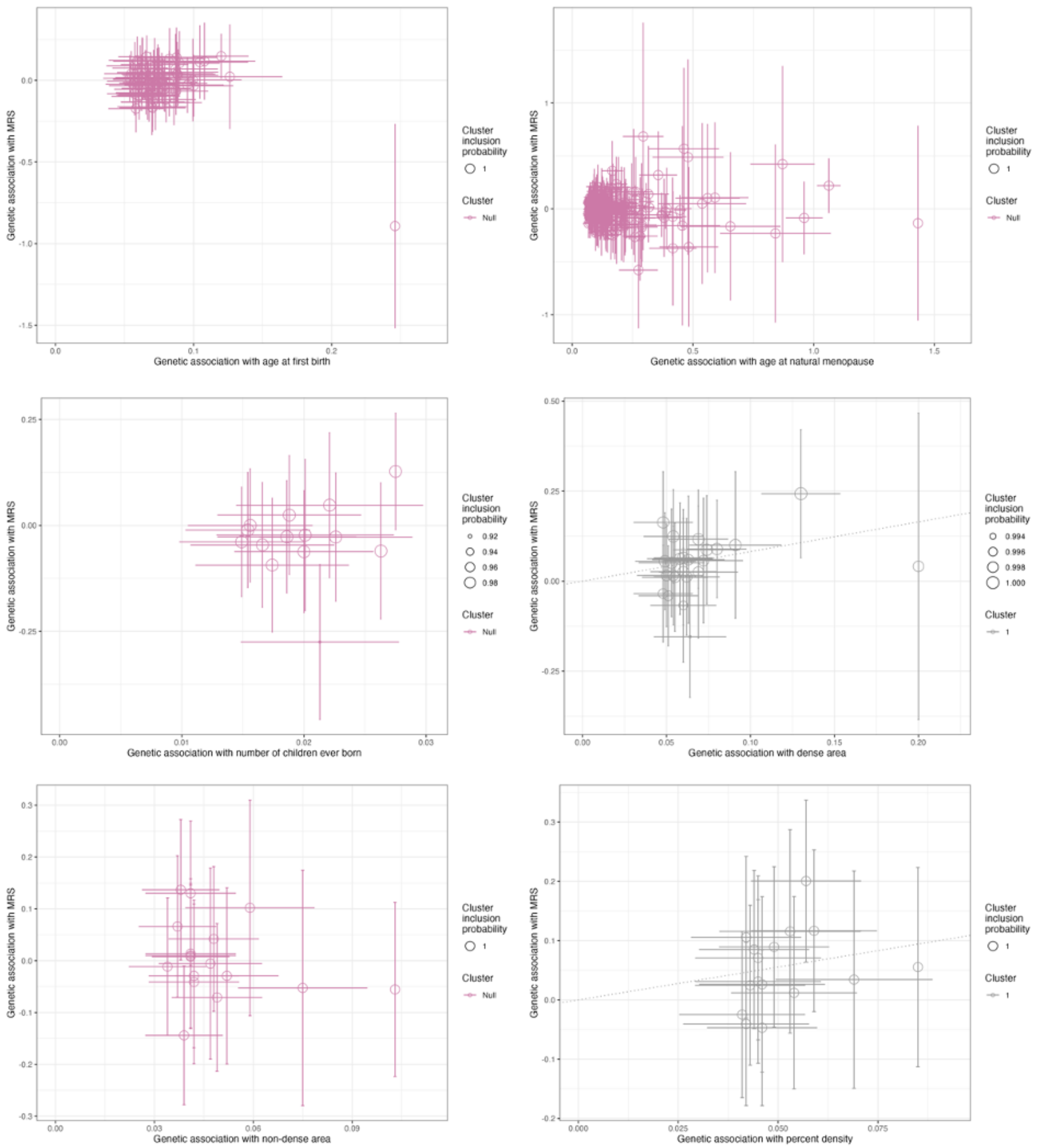


**Figure S3 (continued).** Genetic associations between each risk factor and mammogram risk score (MRS). Scatter plots show results from MR-Clust analysis, with genetic association with MRS (y-axis) versus genetic association with each risk factor (x-axis). Each point represents a genetic variant, with lines indicating 95% confidence intervals.

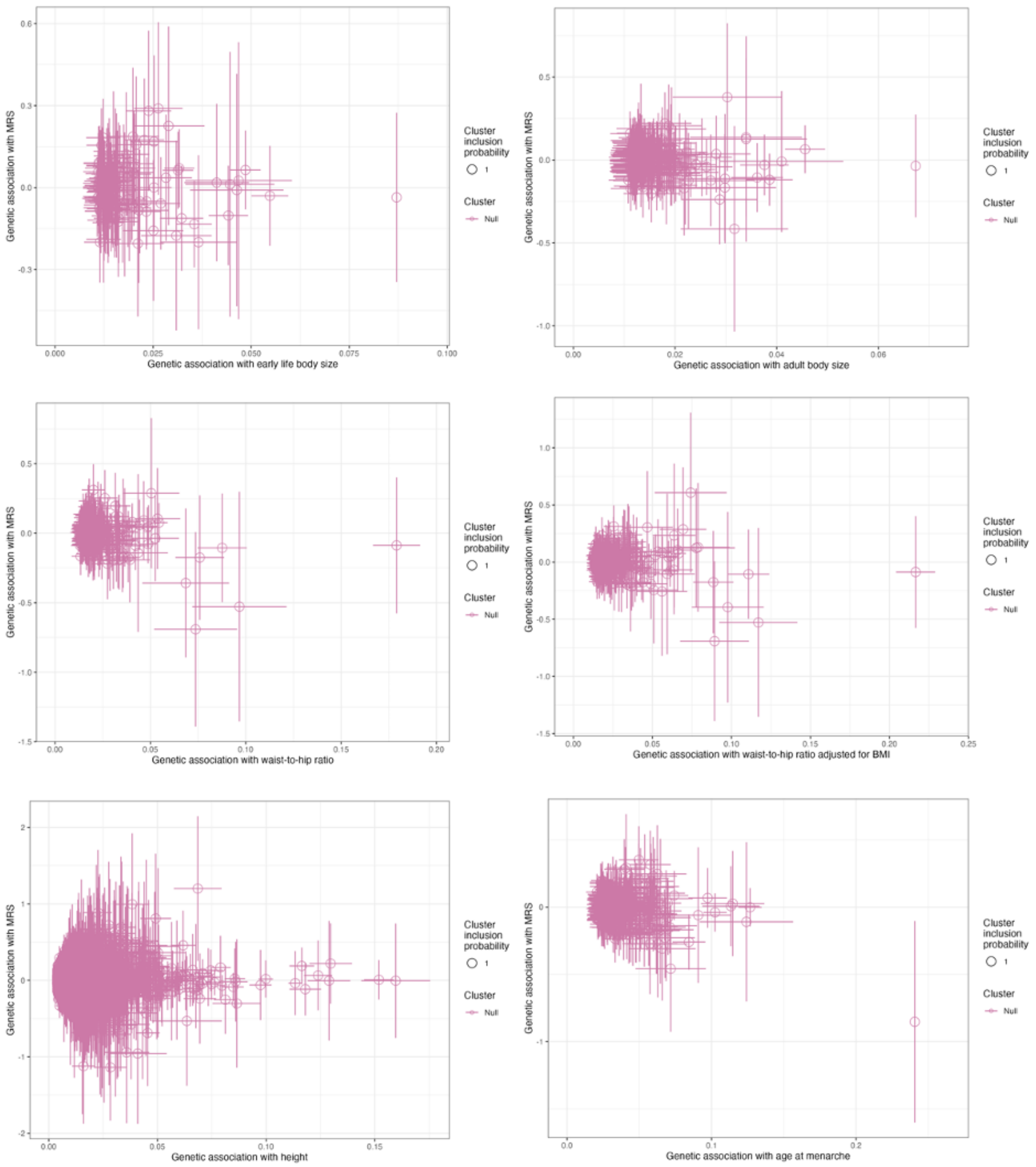


**Figure S4.** Genetic associations between each risk factor and mammogram risk score (MRS) using instrumental variable-MRS associations adjusted for menopausal status. Scatter plots show results from MR-Clust analysis, with genetic association with MRS (y-axis) versus genetic association with each risk factor (x-axis). Each point represents a genetic variant, with lines indicating 95% confidence intervals.

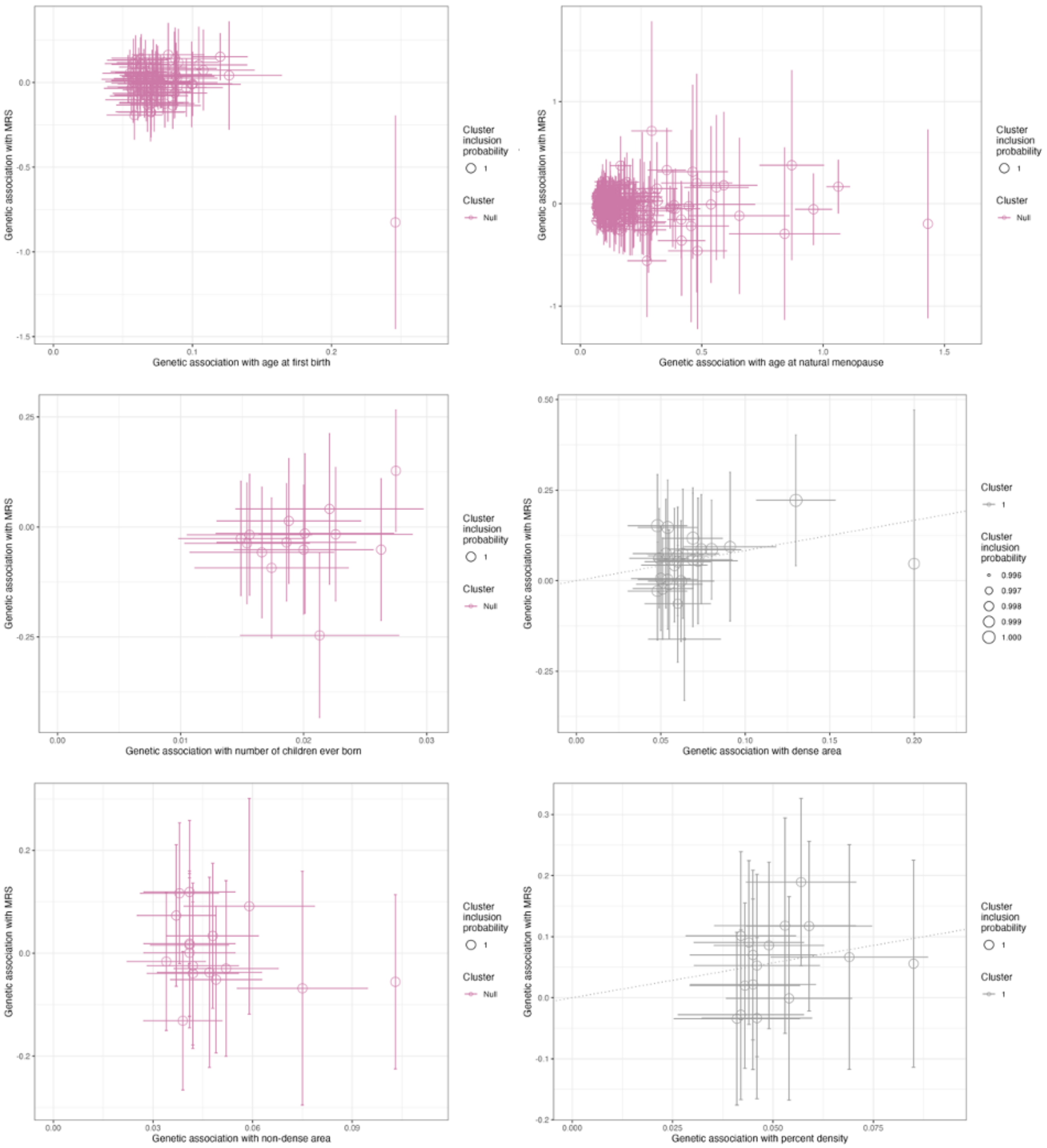


**Figure S4 (continued).** Genetic associations between each risk factor and mammogram risk score (MRS) using instrumental variable-MRS associations adjusted for menopausal status. Scatter plots show results from MR-Clust analysis, with genetic association with MRS (y-axis) versus genetic association with each risk factor (x-axis). Each point represents a genetic variant, with lines indicating 95% confidence intervals.

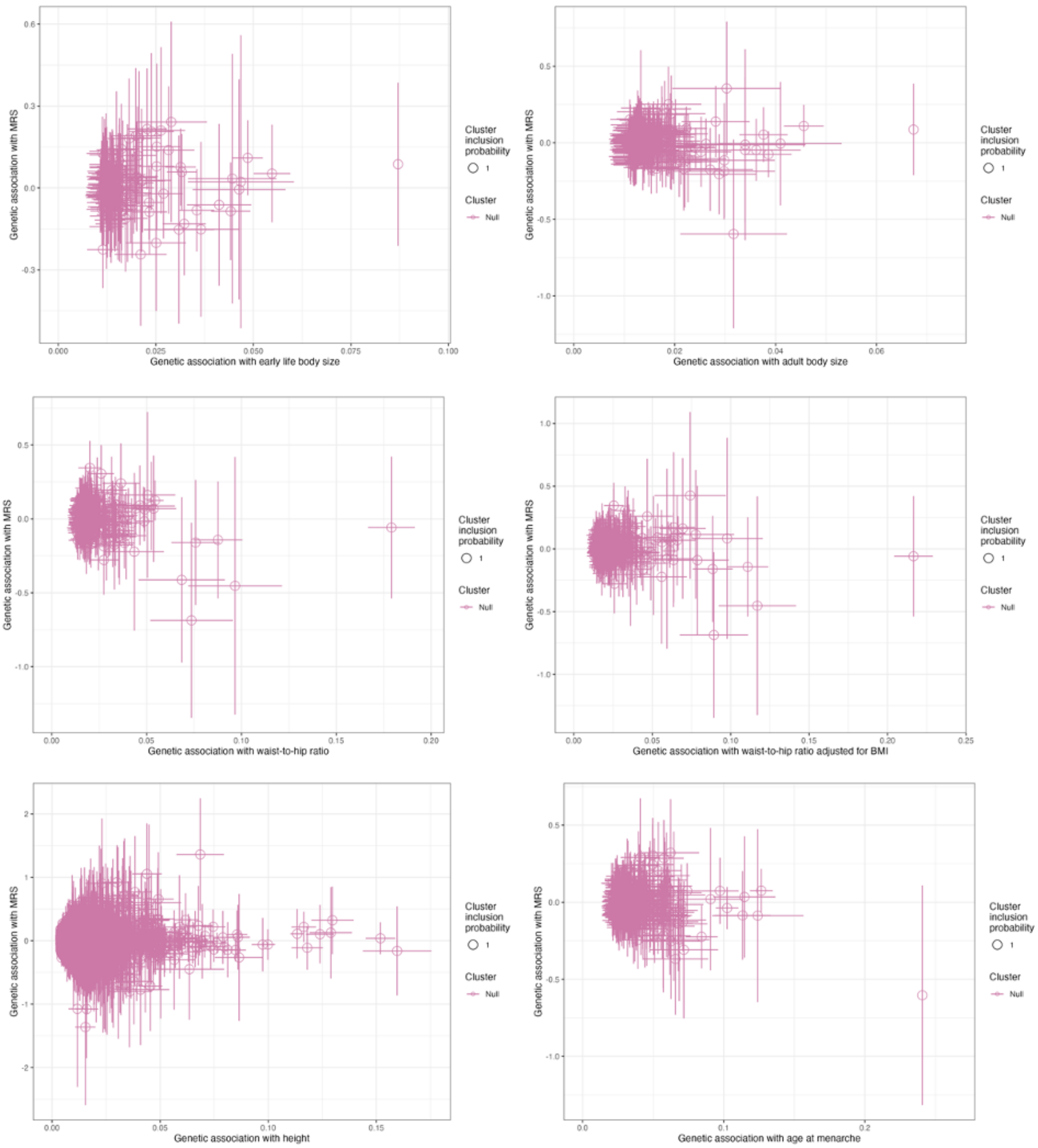


**Figure S5.** Genetic associations between each risk factor and mammogram risk score (MRS) using instrumental variable-MRS associations adjusted for predicted BI-RADS density. Scatter plots show results from MR-Clust analysis, with genetic association with MRS (y-axis) versus genetic association with each risk factor (x-axis). Each point represents a genetic variant, with lines indicating 95% confidence intervals.

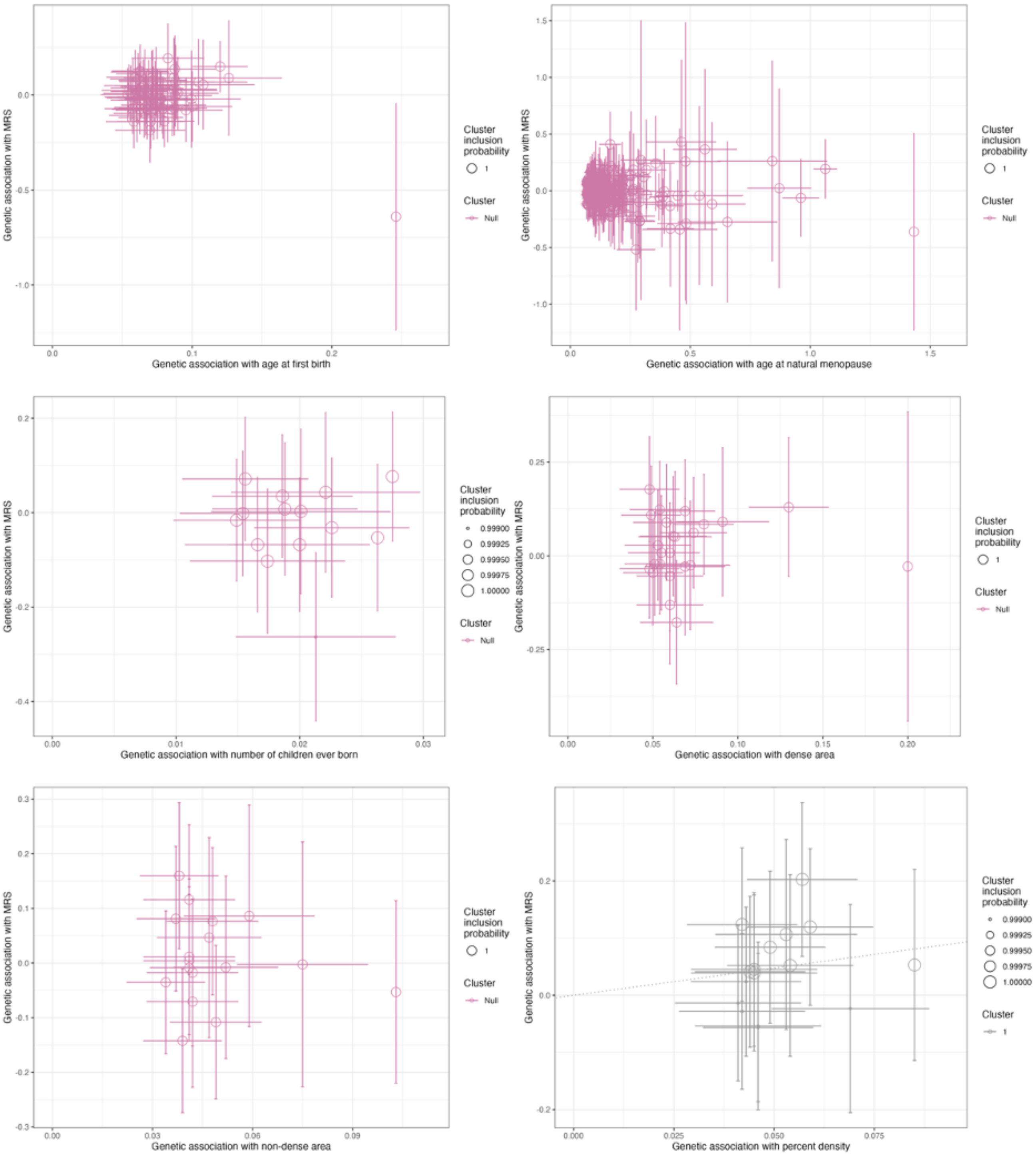


**Figure S5 (continued).** Genetic associations between each risk factor and mammogram risk score (MRS) using instrumental variable-MRS associations adjusted for predicted BI-RADS density. Scatter plots show results from MR-Clust analysis, with genetic association with MRS (y-axis) versus genetic association with each risk factor (x-axis). Each point represents a genetic variant, with lines indicating 95% confidence intervals.
